# Impact of Azithromycin Administration at Hospital Discharge on Antimicrobial Resistance and Enteropathogen Carriage 3 Months Following Treatment

**DOI:** 10.64898/2026.04.17.26351054

**Authors:** Polycarp Mogeni, John Benjamin Ochieng, Kevin Kariuki, Doreen Rwigi, Hannah E. Atlas, Kirkby D. Tickell, Laura Riziki Aluoch, Catherine Sonye, Evans Apondi, Lilian Ambila, Mame M. Diakhate, Benson O. Singa, Jie Liu, James A. Platts-Mills, Queen Saidi, Donna M. Denno, Ferric C. Fang, Judd L. Walson, Eric R. Houpt, Patricia B. Pavlinac

**Affiliations:** Center for Microbiology Research, Kenya Medical Research Institute (KEMRI), Nairobi, Kenya; Department of Global Health, University of Washington, Seattle, Washington, United States of America; Center for Global Health Research, Kenya Medical Research Institute (KEMRI), Kisumu, Kenya; Center for Clinical Research, Kenya Medical Research Institute (KEMRI), Nairobi, Kenya; School of Public Health, Qingdao University, Qingdao, China; Division of Infectious Diseases and International Health, Department of Medicine, University of Virginia, Charlottesville, Virginia, USA; Department of Pediatrics, University of Washington, Seattle, Washington, United States of America; Department of Medicine, Division of Allergy and Infectious Diseases, University of Washington, Seattle, Washington, United States of America; Department of Laboratory Medicine and Pathology, University of Washington, Seattle, Washington, United States of America; Department of Microbiology, University of Washington, Seattle, Washington, USA; The Childhood Acute Illness & Nutrition (CHAIN) Network, Nairobi, Kenya; Departments of International Health, Johns Hopkins University, Baltimore, Maryland, United States of America; Department of Medicine, Johns Hopkins University, Baltimore, Maryland, United States of America; Department of Pediatrics, Johns Hopkins University, Baltimore, Maryland, United States of America; Department of Epidemiology, University of Washington, Seattle, Washington, United States of America

**Keywords:** AMR, macrolide resistance, azithromycin, enteric pathogens, post-discharge

## Abstract

**Background:** The Toto Bora trial tested whether a course of azithromycin reduced rates of re-hospitalization or death in the 6 months following hospitalization among Kenyan children. We hypothesized that azithromycin would reduce enteric bacteria and increase carriage of macrolide resistance in the subsequent 3 months.

**Methods:** Kenyan children (1-59 months) hospitalized and subsequently discharged for non-traumatic conditions provided fecal samples before and 3 months after randomization to a 5-day course of azithromycin or placebo. Quantitative PCR identified enteropathogens and AMR-conferring genes in fecal samples. Generalized estimating equations assessed the impact of the randomization arm on pathogen and resistance gene detection, accounting for baseline presence and site.

**Results:** Among 1,393 baseline stools, 12.4% had at least one bacterial enteropathogen, 94.7% had at least one macrolide-resistance gene, and 92.6% had at least one beta-lactamase-resistance gene identified. At month 3, children randomized to azithromycin had a 6.1% higher likelihood of carrying a macrolide resistance gene compared to placebo (adjusted prevalence ratio [aPR], 1.06; 95% CI, 1.04–1.08; P<0.001). Specifically, azithromycin randomization was associated with a higher relative prevalence of *erm(B)* (aPR, 1.09 [95% CI, 1.04-1.15]; P=0.001), *erm(C)* (aPR, 1.23 [95% CI, 1.14-1.31]; P<0.001), *msr(A)* (aPR, 1.14 [95% CI, 1.04-1.25]; P=0.007), and *msr(D)* (aPR, 1.07 [95% CI, 1.03-1.11]; P=0.001). There was no difference in overall bacterial pathogen prevalence (18.9% vs 17.3%) between randomization arms, but a slightly lower proportion of children had *Shigella* after randomization in the azithromycin arm (3% vs. 5%, aPR, 0.79 [95% CI, 0.62, 1.01]; P=0.063).

**Interpretation:** Azithromycin at hospital discharge was associated with higher carriage of macrolide-resistance-conferring genes in the post-discharge period compared with placebo, without significant declines in enteric pathogen carriage other than modest changes to *Shigella*. The potential benefits and risks of empiric azithromycin need to be considered, as children are increasingly exposed to this broad-spectrum antibiotic.

## Introduction

In settings of high childhood mortality, mass drug administration (MDA) of azithromycin is a possible mortality-reducing strategy [1,2] endorsed by the World Health Organization [3]. The mechanisms by which azithromycin MDA reduces mortality are not fully understood, but may include a reduction in the incidence and severity of various infectious diseases [4], including enteric diseases. Azithromycin MDA in Niger, where Shigella is the leading cause of dysentery, was associated with a reduction in *Shigella* fecal carriage [5] and dysentery-related mortality [4]. Azithromycin given to 6-11-month-old infants in India reduced fecal carriage of enteroaggregative (EAEC), enteropathogenic (EPEC), and enterotoxigenic *Escherichia coli* (ETEC), *Campylobacter*, and *Salmonella*, and decreased concentrations of fecal biomarkers of enteric inflammation 2 weeks later [6]. Pathogenic enteric bacteria may underlie the empiric mortality benefit associated with azithromycin.

The potential benefits of azithromycin must be carefully weighed against its risks, including induction of antimicrobial resistance [7]. Studies of azithromycin MDA have documented increased macrolide-resistant *Streptococcus pneumoniae* nasopharyngeal carriage [8–10] and selection pressure in the gut microbiota, resulting in increased macrolide-resistant *E coli* in fecal samples [7,11]. Furthermore, a recent MDA study observed increased resistance to antimicrobial classes beyond macrolides, indicating the potential for broader selection of resistance following azithromycin MDA [12]. This unintended consequence poses a substantial threat, potentially undermining the long-term effectiveness of azithromycin and other vital antibiotics, complicating future efforts to control infectious diseases.

The Toto Bora trial was an individual randomized placebo-controlled trial that tested whether a 5-day course of azithromycin delivered at hospital discharge prevented hospitalization and death in Kenyan children [13]. The trial found no reduction in death or hospitalization [14], consistent with the lack of mortality benefit observed in a recent individually randomized trial of azithromycin delivered at routine infant health visits [15]. We previously reported high antimicrobial resistance in *E coli* [16,17] and *Klebsiella* spp. [18] isolates from a subset of children at baseline in the Tota Bora trial. In this cohort, the clinical benefit of azithromycin was dependent on the presence of the *mef(A)* resistance gene: azithromycin reduced the hazard of death or rehospitalization by 34% in children without the gene, yet was associated with a more than two-fold increase in risk among those carrying the gene.

While these prior analyses focused on baseline isolate resistance and clinical outcomes, the current study investigates the longitudinal impact of treatment on the gut environment. We hypothesized that azithromycin would reduce the prevalence of enteric bacterial pathogens while concurrently driving the expansion of the fecal resistome, rather than just resistance in individual isolates, during the three months following administration.

## Methods

### Ethical Statement

The KEMRI Scientific and Ethics Review Unit (KEMRI; SERU 3086, Sept 8, 2015), the Kenya Pharmacy and Poisons Board (ECCT/15/10/04, Dec 3, 2015), and the University of Washington (IRB 49120, June 2, 2015) approved the study. Caregivers provided written informed consent for study participation and sample provision for antimicrobial resistance testing. The trial, registered on ClinicalTrials.gov (NCT02414399), adhered to the principles outlined in the International Conference on Harmonization Good Clinical Practice, the Declaration of Helsinki, and the ethical requirements in Kenya.

### Study Design, Intervention, and Sample Collection

A detailed description of the Toto Bora trial has been published elsewhere [13]. This was a two-arm, parallel-group, randomized controlled trial (RCT) conducted from 2016 to 2019 across four Kenyan hospitals: Kisii Teaching and Referral, Homa Bay Teaching and Referral, St. Paul Mission, and Kendu Adventist Mission Hospitals. The study evaluated the impact of azithromycin, administered at hospital discharge, on the risk of death and rehospitalization among children aged 1-59 months. Participants were randomized 1:1 to receive a 5-day course of either oral azithromycin suspension (10 mg/kg on Day 1, followed by 5 mg/kg on Days 2–5) or an identically formulated placebo. The first dose was directly observed at the hospital, while subsequent doses were administered by caregivers at home. Children were ineligible if they were admitted for trauma, poisoning, or a congenital anomaly [13]. In addition, the study also excluded those who had a documented macrolide allergy, were prescribed a macrolide antibiotic at discharge, or were taking a protease inhibitor for HIV infection. Finally, children were excluded if their caregiver was not the legal guardian or did not intend to remain in the study area for at least 6 months, or if a same-sex twin had already been enrolled [13]. Stool or rectal swab samples were collected at enrollment and three months post-randomization, and promptly stored at -80°C to preserve sample integrity.

### Laboratory Analysis

Detailed laboratory methods are described elsewhere [19]. In summary, between 2022 and 2023, enrollment and month-3 stool/rectal swab samples were subjected to qPCR at KEMRI-CGHR in Kisumu. DNA was extracted using the QIAamp Fast Stool Mini Kit, modified for bead beating and heat incubation. TaqMan array cards targeting prespecified enteropathogens and antibiotic resistance genes were used (Supplementary Table 1), enabling simultaneous detection of a wide range of pathogens and resistance markers in a single assay. Bacteriophage MS2 and phocine herpesvirus served as extraction and amplification controls. qPCR was performed on the ViiA7 platform (Applied Biosystems, Thermo Scientific), and target amplification cycle threshold (Ct) values were securely recorded in a password-protected database (The Multi-Schema Information Capture [MuSIC] database) jointly managed by the University of Virginia and the collaborating local laboratory.

### Outcome

The primary prespecified outcome was the presence of macrolide resistance determinants: *erm(A), erm(B), erm(C), mef(A), mph(A), mph(B), msr(A)*, and *msr(D)*, or leading bacterial causes of diarrhea [20]: Typical enteropathogenic *Escherichia coli* (tEPEC), Heat-stable toxin-producing enterotoxigenic *E. coli* (ST-ETEC), *Shigella*, and *Campylobacter jejuni/coli*. Secondary outcomes included enteric bacteria not included in the primary outcomes, enteric viruses and protozoa (Supplementary Table 1), and beta-lactam resistance-conferring genes (Supplementary Table 1). AMR gene and enteric pathogen positivity were determined using a Ct <30 to exclude low gene quantity, and sensitivity analysis was conducted using a Ct <35 cutoff.

### Statistical analyses

We used a generalized estimating equations (GEE) model with a modified Poisson regression model to compare the proportions of enteric bacteria and macrolide resistance genes detected between the azithromycin and placebo arms. Subgroup analyses included participants hospitalized for gastroenteritis. Models were adjusted for baseline detection of the pathogen or AMR gene of interest, as well as study site. All statistical analyses were performed using two-sided tests. A Bonferroni-adjusted significance threshold of P<0.007 for macrolide-resistance genes and P<0.013 for bacterial pathogens was applied to account for multiple comparisons in the primary outcome analyses, including both main and subgroup analyses. Exploratory analyses also assessed additional pathogens and antibiotic resistance genes present in over 2% of baseline fecal samples. All statistical analyses were performed using R version 4.4.1 and Stata version 18 (StataCorp) software.

## Results

The Toto Bora trial recruited 1,400 children at hospital discharge who were randomly assigned to receive either azithromycin or a placebo. A detailed description of the baseline study population has been reported previously [14]. In brief, two children were withdrawn due to ineligibility, five had insufficient baseline samples, 22 died before month 3, and 135 had no month 3 sample because they did not return for follow-up or had moved out of the study area or had insufficient sample for processing (Figure 1). Of the 1236 children included in this analysis, 628 (50.8%) received azithromycin, and 608 (49.2%) received a placebo (Figure 1). At baseline, the median age was 18 months (interquartile range [IQR], 9–32 months); 508 children (41.1%) were female, and 114 (9.2%) were wasted. Most children (n = 1,104, 89.3%) received antibiotic treatment during their hospital stay before randomization, of whom 3.7% received a macrolide antibiotic (Table 1).

**Figure 1.**
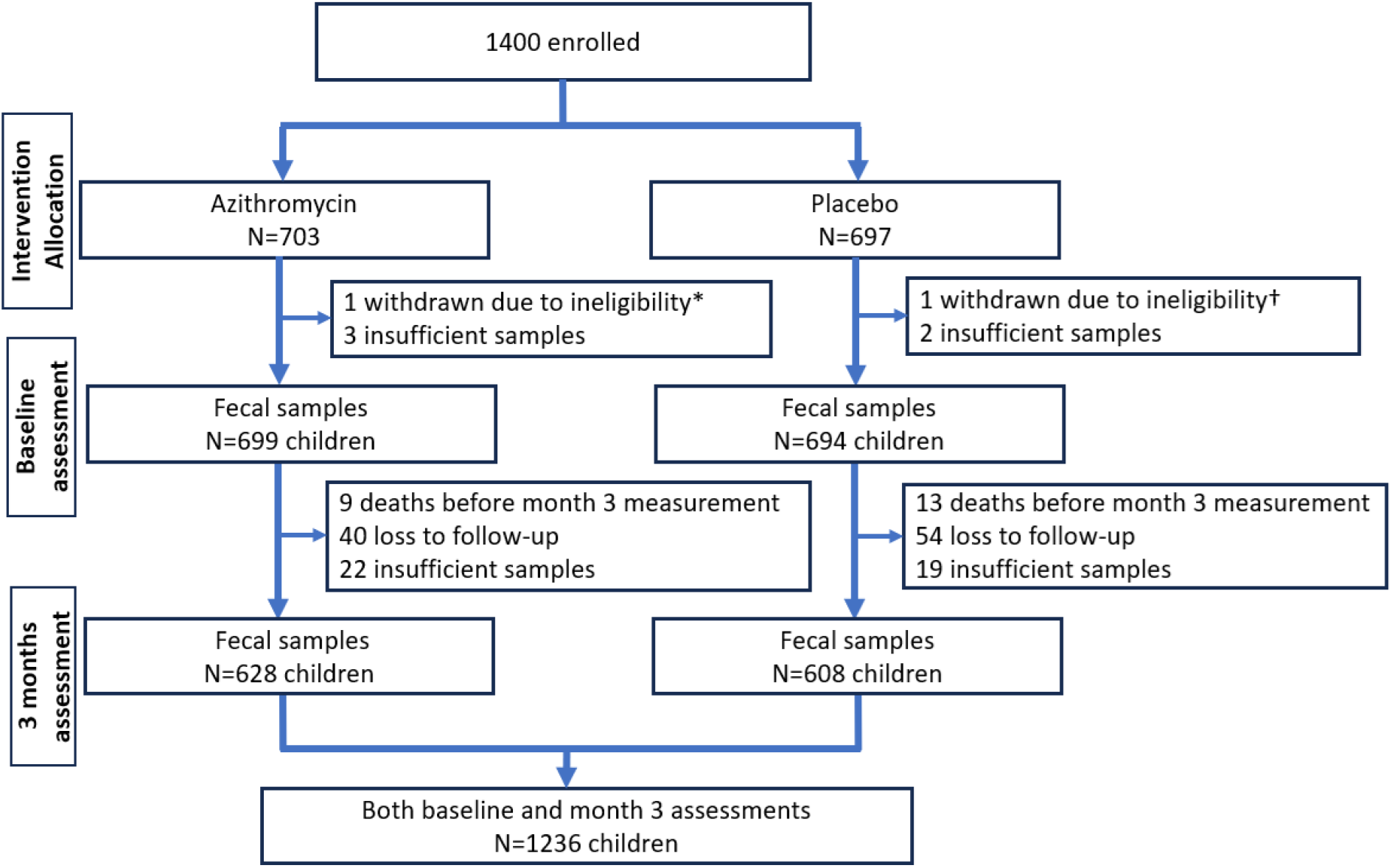
Study profile. *An admission for poisoning alone, which is an exclusion criterion, and a †twin whose sibling had already been enrolled.

**Table 1.**
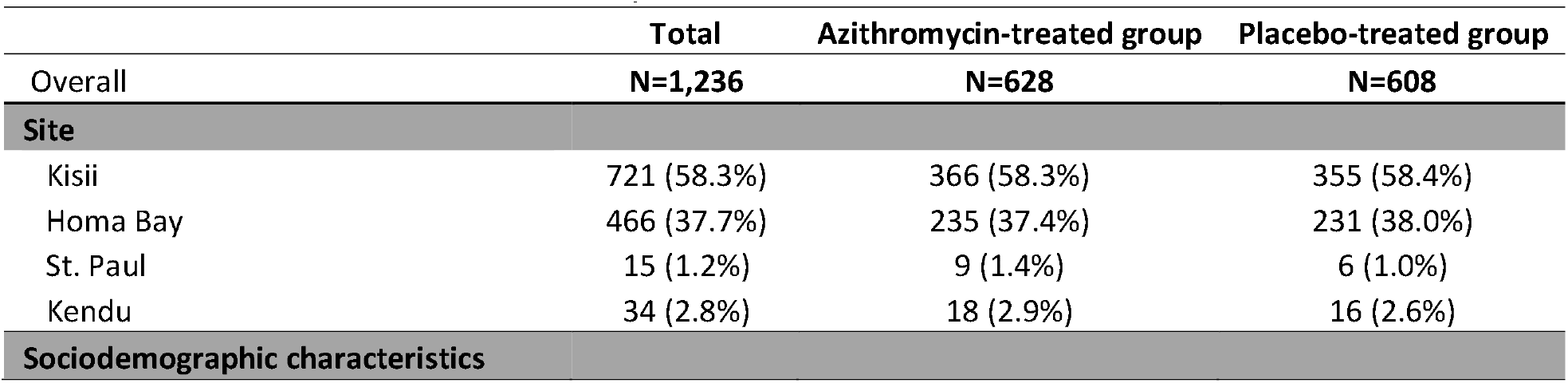
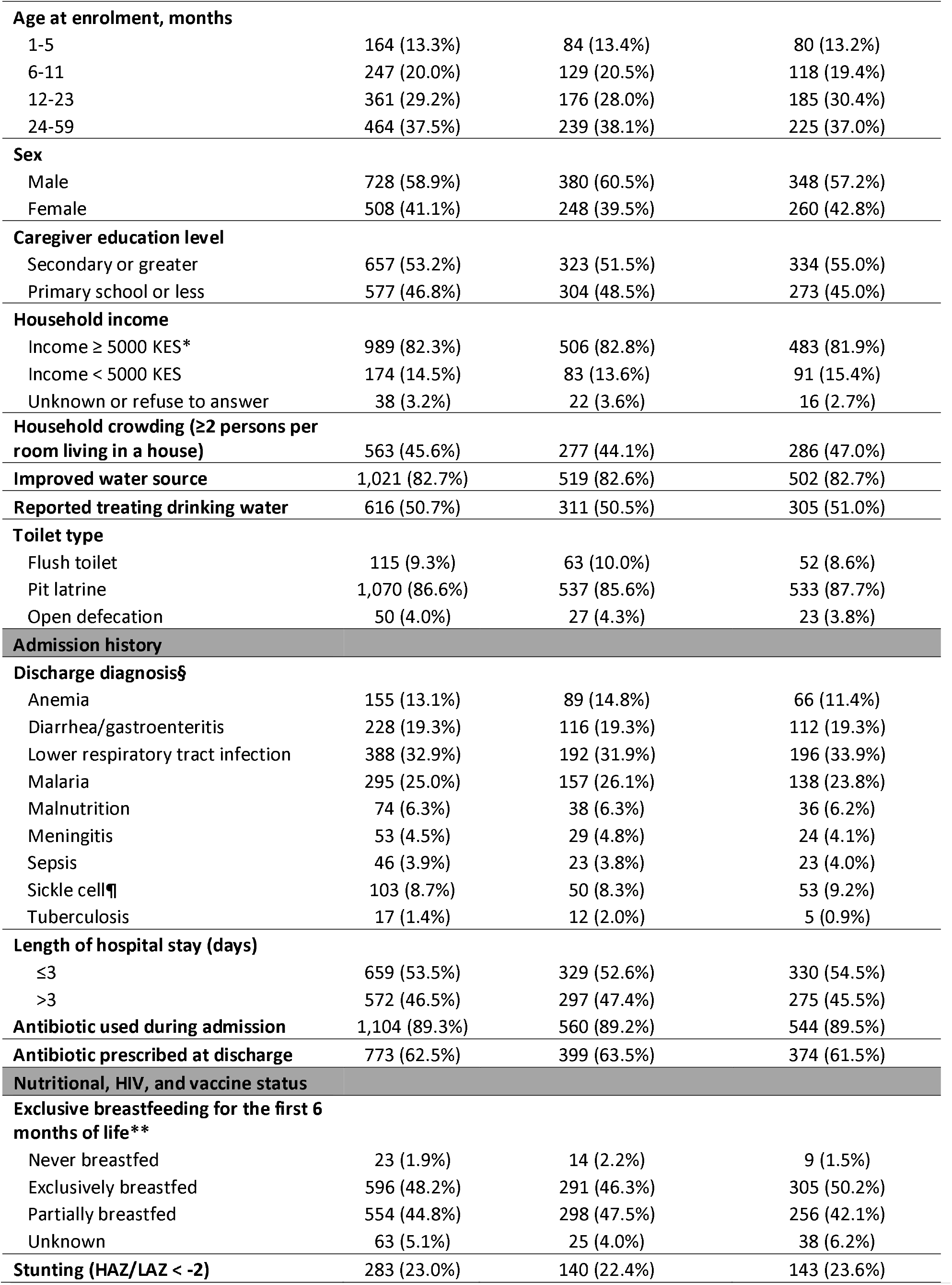
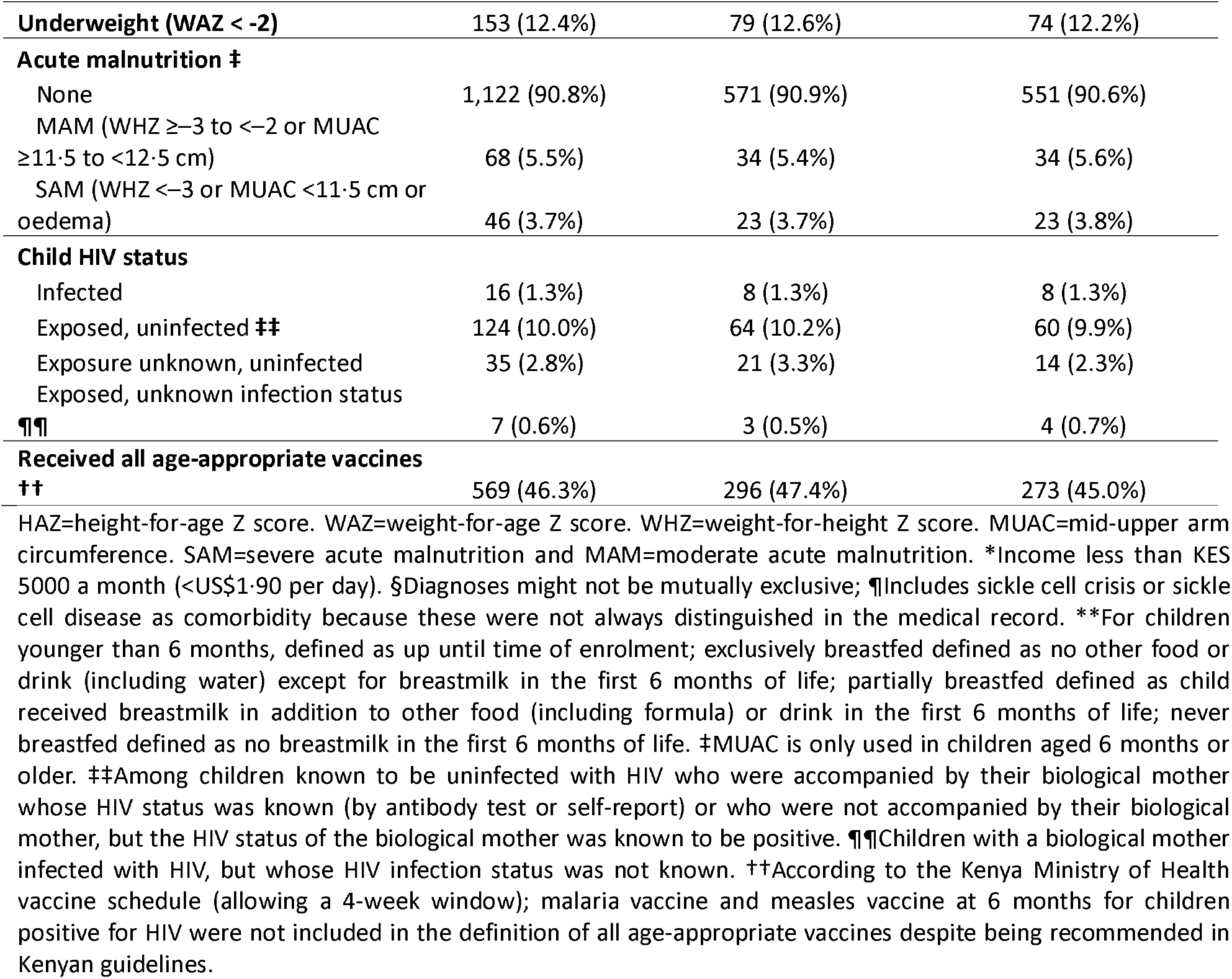
Baseline characteristics overall and by randomization group among children with both baseline and month 3 fecal sample TAC results.

Most fecal samples were from rectal swabs (579 [92.2%] and 559 [91.9%] in azithromycin and placebo arms, respectively, at baseline and 628 [100%] and 608 [100%] in azithromycin and placebo arms, respectively, at month 3); the remaining samples were from whole stool. At baseline, 94.7% of fecal DNA samples contained macrolide resistance genes, decreasing to 86.1% (χ^2^ -test; P<0.001) at month 3 (Figure 2 and in more detail in Supplementary Table 1). The prevalence of individual genes changed as follows: *erm(A)*, 15.6% to 9.7%; *erm(B)*, 60.7% to 40.9%; *erm(C)*, 41.3% to 28.9%; *mef(A)*, 23.7% to 23.0%; *mph(A)*, 68.6% to 40.3%; *msr(A)*, 22.3% to 6.9%; and *msr(D)*, 67.3% to 63.3%. In models adjusted for baseline detection and site, children receiving azithromycin had a 6.1% (adjusted prevalence ratio (aPR), 1.06 [95% CI, 1.04-1.08]; P<0.001) higher likelihood of carrying a macrolide gene in their fecal sample 3 months after randomization compared with children in the placebo arm (Figure 3). Azithromycin was specifically associated with higher prevalences of *erm(B)* (aPR, 1.09 [95% CI, 1.04-1.15]; P=0.001), *erm(C)* (aPR, 1.23 [95% CI, 1.14-1.31]; P<0.001), *msr(A)* (aPR, 1.14 [95% CI, 1.04-1.25]; P=0.007), and *msr(D)* (aPR, 1.07 [95% CI, 1.03-1.11]; P=0.001) at month 3 (Figure 3). Although there was a relatively higher prevalence of *mef(A)* (aPR, 1.12 [95% CI, 1.02-1.23]; P=0.021) among children receiving azithromycin, this was not statistically significant after Bonferroni adjustment for multiple testing. These findings were also observed in a sensitivity analysis using CT<35 (Supplementary Figure 1).

**Figure 2:**
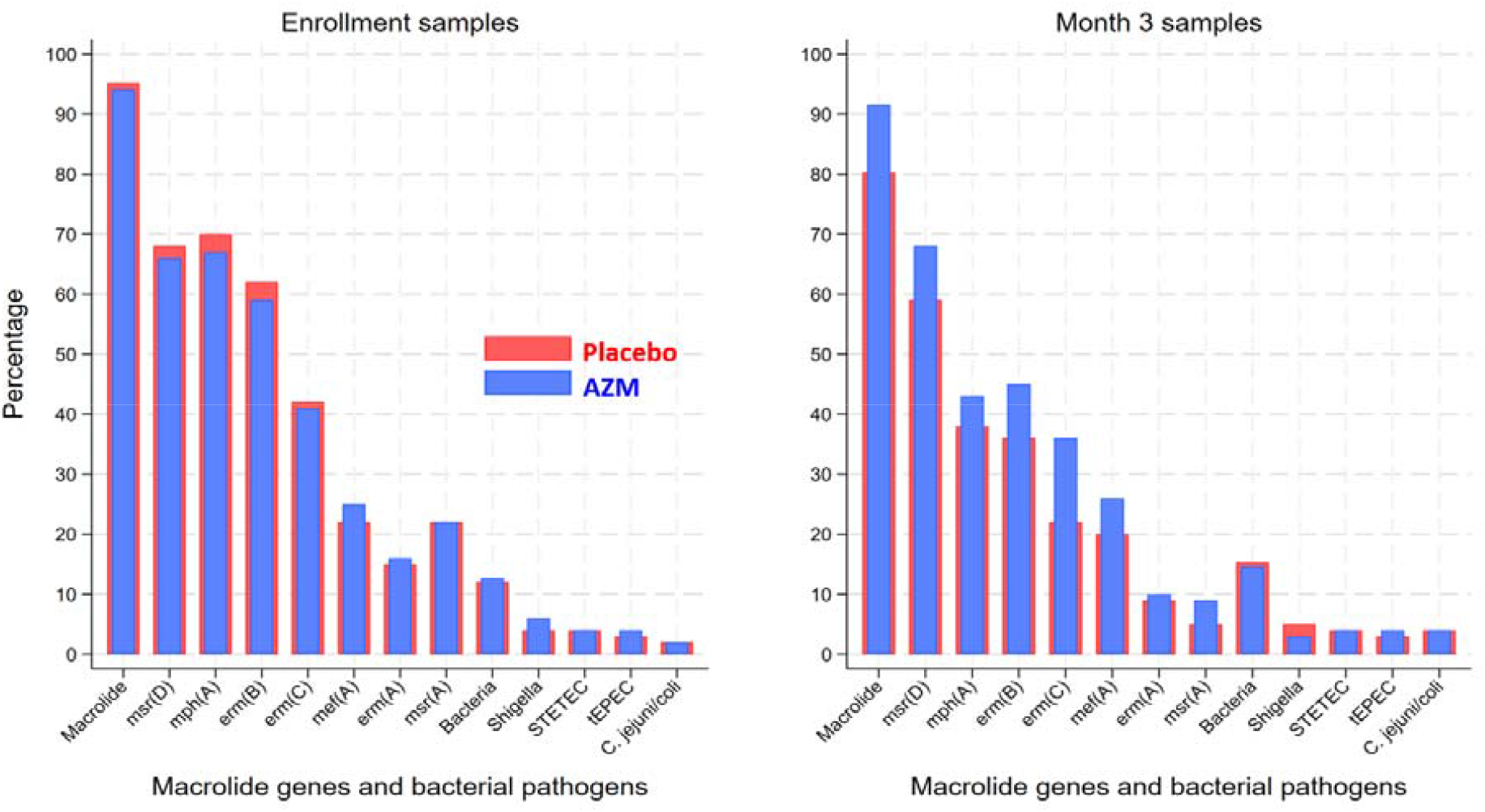
Percentage of macrolide resistance gene and pathogenic bacteria carriage in fecal samples from children at hospital discharge and 3 months post-discharge. Percentages are presented by randomization arm at baseline and three months following randomization to azithromycin and placebo. Bacteria refers to the presence of any of the following pathogens: typical enteropathogenic *E. coli* (tEPEC), heat-stable toxin-producing enterotoxigenic *E. coli* (ST-ETEC), *Shigella*, and *Campylobacter jejuni/coli*.

**Figure 3:**
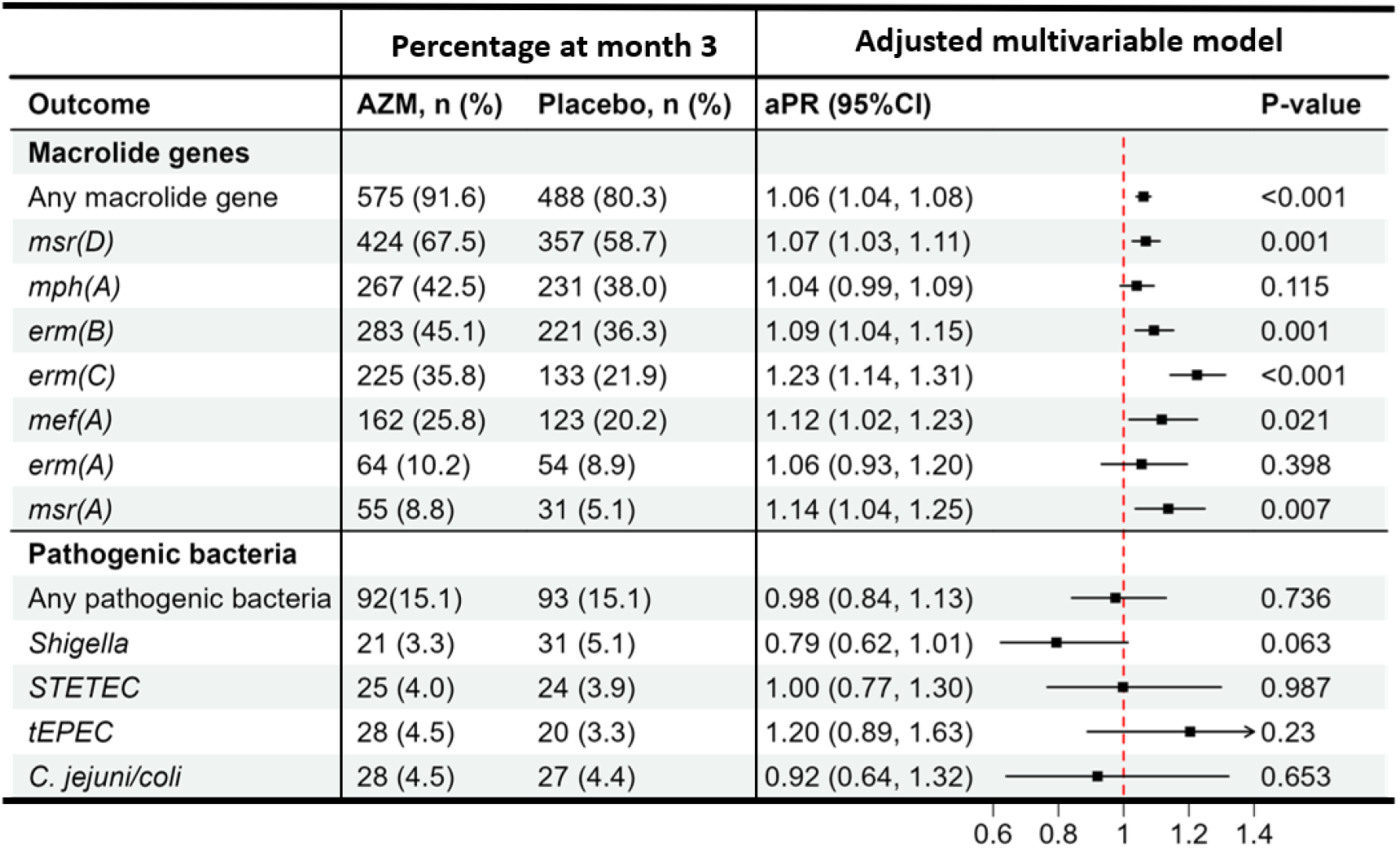
Azithromycin’s Effect on 3-Month Detection of Macrolide Resistance Genes and Pathogenic Bacteria (cycle threshold (CT) <30). Black square dots and error bars represent adjusted prevalence ratios and 95% confidence intervals, respectively, illustrating the impact of azithromycin on macrolide resistance genes and pathogenic bacteria (defined by CT<30) in fecal DNA from discharged children. *Vibrio cholerae* and *Salmonella* were excluded from the primary bacterial pathogens due to infrequent detections in the dataset. Each GEE with a modified Poisson regression model was adjusted for baseline detection and study site. Multiple comparisons were accounted for using a Bonferroni-adjusted significance level of <0.007 for macrolide genes and <0.013 for bacterial pathogens. “Any pathogenic bacteria” refers to the presence of any of the following pathogens: typical enteropathogenic *E. coli* (tEPEC), heat-stable toxin-producing enterotoxigenic *E. coli* (ST-ETEC), *Shigella*, and *Campylobacter jejuni/coli*.

At baseline, 12.4% of samples contained at least one of the four relevant diarrhea-causing bacterial enteric pathogens, compared to 15.0% at month 3, with no difference between arms (aPR, 0.98 [95% CI, 0.84-1.13]; p=0.736) (Figure 2 and Supplementary Table 1). Specific pathogen detections tested at baseline and month 3, respectively, were: *Shigella* (4.7% and 4.2%), tEPEC (3.2% and 3.9%), STETEC (4.0% and 3.9%), and *Campylobacter jejuni or coli*. (2.0% and 4.5%). In multivariable models adjusted for baseline pathogens, there was no difference in the proportion of pathogenic bacteria between randomization arms at month 3 (aPR, 0.98 [95% CI, 0.84-1.13]; P<0.736). In addition, none of the individual primary pathogenic bacteria were impacted by azithromycin at the three-month timepoint, although *Shigella* trended toward a lower prevalence in the azithromycin arm (aPR, 0.79 [95% CI, 0.62-1.01]; P=0.063) (Figure 3), which was not statistically significant. These findings were consistent in the sensitivity analyses based on Ct<35 (Supplementary Figure 1). In a subgroup analysis among 228 children who had been hospitalized with a diagnosis of diarrhea (116 azithromycin and 112 placebo), azithromycin was associated with a lower baseline-adjusted prevalence of *Shigella* detection at 3 months (aPR, 0.47 [95% CI, 0.25-0.88]; P=0.018), but not with reductions in other bacterial enteropathogens (Supplementary Figure 2). Enteropathogen Ct values tended to be higher at discharge in children who were admitted for diarrhea than in those without (Supplementary Table 2).

In exploratory analyses, we evaluated the impact of azithromycin on the other enteric bacterial pathogens, viruses, protozoa, and beta-lactam resistance-conferring genes on the TAC platform. Enteric viruses were detected in 15.6% of samples at baseline and in 6.0% at month 3, a reduction that was particularly notable for cytomegalovirus (CMV, 5.3% and 1.4%, respectively). In contrast, changes in other viruses, protozoa, and parasites were modest (Supplementary Table 1). CMV was present in 0.6% of month-3 samples from children randomized to azithromycin and 2.3% in those randomized to placebo (aPR, 0.79 [95% CI, 0.66-0.95]; p=0.012). The prevalence of samples with at least one beta-lactamase-resistance gene also decreased, from 92.6% at baseline to 89.8% at month 3 (Supplementary Table 1). Both in the analyses including all children (Supplementary Figure 3 and Supplementary Figure 4) and in the gastroenteritis subgroup analyses (Supplementary Figure 5), there was no evidence of azithromycin’s effect on beta-lactamase genes.

## Discussion

Although azithromycin has demonstrated survival benefits in some high-risk pediatric populations, likely in part through the reduction of bacterial pathogens in the gut [1,5], its potential to promote antimicrobial resistance raises important concerns for clinical and public health practice [21]. In this nested analysis of an individually randomized trial of children recently discharged from health facilities in western Kenya, a 5-day course of azithromycin was associated with slightly higher macrolide resistance gene carriage at three months compared with placebo. The high carriage of macrolide-resistance genes in the gut of discharged children reduced after three months, and to a greater extent among children randomized to placebo versus azithromycin. While there was no significant difference in the overall prevalence of enteric bacterial pathogens, other than a trend for *Shigella*, in the subset of children hospitalized for diarrhea, a population with the likely highest quantity of *Shigella* infections, azithromycin did appear to reduce Shigella carriage 3 months later. Taken together, these findings substantiate evidence that azithromycin may reduce *Shigella* carriage among children with diarrhea, but also highlight the risk azithromycin poses in promoting carriage of potentially transmissible macrolide-resistance genes during the post-discharge period.

Pathogen-specific analyses suggested a reduction in *Shigella* among children who received azithromycin; however, this effect was greatest in children hospitalized for diarrhea, the population most likely to have *Shigella* at the time of azithromycin treatment. Azithromycin given to Bangladeshi children with susceptible *Shigella* diarrhea had a reduction in *Shigella* carriage 5-7 days later [22]. The median duration of *Shigella* carriage following diarrhea is 14 days [23], and our findings suggest that Shigella carriage may persist for at least 3 months.

Treating *Shigella* diarrhea with azithromycin has shown clinical and growth benefits [20], and reduced carriage may also translate to reduced transmission. The detection of *Shigella* in this discharged population of children hospitalized for all causes was low, at less than 5% at baseline, which contrasts with the high prevalence observed in the MDA trial in Niger (14.1%) where MDA azithromycin had a mortality benefit. If *Shigella* carriage explains a part of azithromycin’s mortality benefit in high-mortality settings, a prevalence threshold above which empiric azithromycin use could be considered might be a plausible targeting strategy.

Azithromycin exposure has been shown to produce transient effects on the gut microbiota and antimicrobial resistance in bacteria that diminish over time [24,25]. For instance, a study of preschool children found that a single dose of azithromycin led to a rapid decrease in microbiome diversity and relatively higher macrolide resistance determinants at two weeks, changes that were no longer detectable at six months [24]. Similarly, in another individually randomized trial, *S. pneumoniae* isolates from school children who received azithromycin showed elevated resistance to erythromycin, oxacillin, and clindamycin at 14 days, with no difference compared to placebo by six months [26]. In addition, our previously published analysis from the Toto Bora trial showed no significant effect of azithromycin on pneumococcal carriage or antimicrobial resistance at month 3 [27], and the ABCD trial similarly found no evidence of phenotypic macrolide resistance at 3- or 6-month follow-up among outpatients with diarrhea randomized to receive azithromycin [28].

Although we observed higher levels of macrolide resistance-conferring genes in fecal samples of children randomized to azithromycin, the clinical relevance of such differences is unclear. With approximately 1200 children (600 per arm) and a baseline proportion of 94% in resistance genes, we were 80% powered to detect differences of ∼5% between arms. A 14% difference (36% vs. 22% in *erm(C)* at Month 3) was the greatest absolute difference we observed, and whether this difference translates to either clinical relevance to individual children and/or indicates greater transmission risk of macrolide-resistant bacteria is not known. Studies of interventions for reducing AMR colonization have found modest to no differences on AMR [29,30].

A recent azithromycin MDA study documented an increase in resistance across antibiotic classes, including in β-lactams, aminoglycosides, metronidazole, trimethoprim, and bacitracin [12]. By contrast, our individually randomized study of azithromycin did not provide evidence of higher β-lactamase genes 3 months later, consistent with a recent individually randomized trial, which showed no evidence of co-selection beyond macrolides [24]. In fact, we hypothesized that azithromycin could reduce carriage of β-lactamase genes if children randomized to azithromycin had fewer infections and fewer antibiotic courses. We observed slightly less β-lactamase carriage, specifically *ACT/MIR* gene carriage, among children randomized to azithromycin, but this was not statistically significant. The absence of major changes in beta-lactam resistance in our trial and in other individually randomized studies of azithromycin [28] suggests that targeted administration may have limited effects on resistance, whereas population-wide exposure through mass drug administration exerts broader and more sustained selection pressures [12].

Our study has important limitations. The three-month sample collection window may have underestimated short-pathogen and resistance-specific effects, as we did not assess carriage at intermediate time points between baseline and three months, nor at the six-month follow-up. This limited our ability to detect transient resistance signals or characterize the rate of change in resistance and gut pathobiome recovery, similar to how a nearly six-month interval after the two-week visit in other studies prevented precise characterization of microbiome and macrolide resistance recovery kinetics. In addition, children who died before the three-month follow-up could not contribute stool samples; because these children were likely at higher risk of harboring resistant organisms, the estimates of resistance presented here may be conservative. Finally, because stool samples reflect only enteric carriage, they may not adequately capture resistance dynamics occurring in other important reservoirs, including the respiratory tract.

From a public health standpoint, our findings underscore the central trade-off in the use of azithromycin among high-risk pediatric populations. Our findings build on an evidence base that *Shigella* carriage is likely reduced by azithromycin, particularly when *Shigella* quantity is high, and that *Shigella* may play a role in explaining azithromycin effects in high mortality settings, which likely overlap with high *Shigella* burden settings. We also add to the mounting evidence that short courses of azithromycin can lead to sustained macrolide-resistance gene expansion; however, the implications for long-term azithromycin susceptibility remain unknown. Any expansion of empiric azithromycin in the context of diarrhea treatment should be embedded within robust resistance surveillance systems and aligned with broader strategies to prevent infection and strengthen child health services.

## Disclaimer

The funder had no role in data collection, analysis, interpretation, or in writing the report or the decision to submit for publication.

## Data Availability Statement

The datasets from the parent Toto Bora trial, linked with the antimicrobial resistance laboratory data utilized in these analyses, will be publicly available on the Harvard Dataverse repository.

## Authors Contributions

**Conceptualization and design:** J.B.O., B.O.S., J.L., J.A.P., F.C.F., J.L.W., E.R.H., P.B.P.

**Performed the experiments:** J.B.O., L.R.A., C.S., E.A., L.A., D.R., K.K., B.O.S., J.L., P.B.P. Data curation: P.M., K.D.T., M.M.D., J.L.

**Funding acquisition:** J.L.W., P.B.P.

**Project administration and investigation:** H.A., K.D.T., M.D.D., B.O.S., Q.S., D.M.D., J.L., P.B.P.

**Formal data analysis:** P.M. Writing original draft: P.M.

**Writing, review and editing:** P.M., J.B.O., H.A., K.D.T., D.R., K.K., L.R.A., C.S., E.A., L.A., M.M.D., B.O.S., J.L., J.A.P., Q.S., D.M.D., F.C.F., J.L.W., E.R.H., P.B.P.

